# Relationship between In-person Instruction and COVID-19 incidence among University Students: A Prospective Cohort Study

**DOI:** 10.1101/2020.08.30.20182139

**Authors:** The Campus & Corona-study Team, Atle Fretheim, Martin Flatø, Arnfinn Helleve, Sølvi Helseth, Gro Jamtvedt, Borghild Løyland, Ida Hellum Sandbekken, Alexander Schjøll, Kjetil Telle, Sara Sofie Viksmoen Watle

## Abstract

Whether university teaching on campus with infection control measures in place is associated with higher risk of COVID-19 than online instruction, is unknown. We will assess this by conducting repeated surveys among students at universities and university colleges in Norway, where some instruction is given in-person, and some is provided online (hybrid model). We will ask about the students’ COVID-19 status, and how much in-person and online instruction they are getting. We will estimate the association between in-person instruction and COVID-19-risk using multivariate regression, controlling for likely confounders. We will also assess whether type of instruction is associated with how satisfied the students are with the instruction, their quality of life, and learning outcomes.

## Background

Social distancing measures, including encouraging people to maintain a distance of 1-2 meters from each other and/or to work from home, are likely to reduce the spread of COVID-19, based on observational evidence and logical arguments [1-3].

Policy makers need to balance the effectiveness of infection control measures against other considerations, including psychosocial and societal consequences. This balance is difficult to strike, especially for institutions where social interactions are of key importance, such as universities. Lack of knowledge about the relative effectiveness of various social distancing measures makes this judgement particularly difficult. As Gressman and Peck put it: “In the absence of relevant prior experience, these institutions are largely in the dark about how one might expect a COVID-19 outbreak to evolve in the unique environment of a college campus and how much of an effect the many possible mitigation strategies should be expected to produce.” [4]

Some attempts have been made at modelling the risk of offering instruction on campus, e.g. a group at Cornell University arrived at the counter intuitive conclusion that shifting to online instruction only would lead to more COVID-19 cases than a full return of students. However, this was premised on “aggressive asymptomatic surveillance where every member of the campus community is tested every 5 days”, as well as sufficient capacity for quarantining, and a series of other assumptions [5].

While universities and colleges in the United States have opted for different models, ranging from completely online to only in-person instruction [6], the higher education institutions in Norway have decided to offer a hybrid model, with some online, and some in-person instruction. This decision enables us to compare students with predominantly in-person instruction and students who receive more online instruction, and to assess the association between campus presence and COVID-19 incidence.

We are not aware of other studies on the association of in-person instruction, or the effect of offering online instruction, on COVID-19 risk. We therefore believe that this study is important to carry out as it has the potential to generate findings that can inform infection control policies at our universities, and in similar environments.

## Methods

We are inviting all universities and university colleges in Norway to take part. All students at institutions that agree to participate will receive an SMS (alternatively an e-mail) inviting them to take part in the study. The invitation includes a link that directs them to a web-based informed consent-form and questionnaire.

We will ask the participants if they have been tested for COVID-19, the results of such a test, how much in-person instruction they have been offered, and how much online instruction they have been offered. We will also inquire about other risk factors for COVID-19 and background variables that may be included as potential adjustment factors (confounders) in the analyses (see Attachment 1 - Questionnaire).

We will survey the students every two weeks by new invitations by SMS or e-mail. The study period will last as long as the universities maintain their arrangements with in-person instruction for only select groups of students. We plan for a study period that lasts for the remainder of 2020.

Students will be asked for consent to link the survey results to information on study programme, basis for admission, study status, academic results, sex and age from the Common Student System

(FS) (see Attachment 2 for full list of variables). Obtaining this information through data linkage will reduce the survey burden for the students and increase the accuracy and quality of the data. The impact of the intervention on exam results and completion will be of significant interest for the institutions and for society at large, as such impacts must be balanced against the needs for disease control. Information on academic performance the previous semester and from upper secondary school will be needed to adjust for these variables as important possible confounders.

We developed a questionnaire through an iterative process, partly based on existing items from existing questionnaires (see Attachment 1 - Questionnaire). Pilot testing with a group of 10 students at Oslo Metropolitan University showed that the questionnaire could be completed with little use of time (around 5 minutes), and that some questions needed to be amended.

We prepared an English version of the questionnaire for students who prefer English over Norwegian. A native English speaker translated the questionnaire to English, and another person not familiar with the questionnaire translated the English version back to Norwegian. We compared the original Norwegian version and the back-translated version, and made minor adjustments.

We have prepared a communication plan, which includes several measures to ensure a high response rate among the students.

Student involvement has taken place at two levels during the project-planning period: We have informed the Student Parliament about the project and they have offered their support, and the pilot testing described above.

### Main outcome

- COVID-19 incidence (self-reported positive test results)

### Secondary outcomes

- Quality of life (“Overall, how satisfied are you with life right now?”)
- Teaching satisfaction (“Overall, how satisfied have you been with the teaching you have received in the past 14 days?”)
- Incidence of COVID-19 testing (self-reported)
- Learning outcomes (from FS – Common Student System)

We will run multivariate regressions to test whether there is an association between in-person instruction and the outcomes.

In-person instruction is a continuous variable, defined as

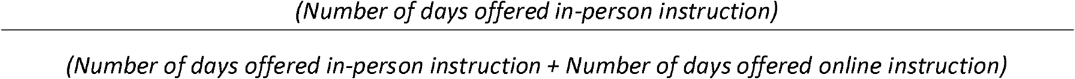

We will run multivariate regressions to test whether there is an association between in-person instruction and the outcomes.

We plan for separate analyses for each participating university/university college, and a pooled analysis across institutions.

### Ethics and data protection

We will use the University of Oslo’s solutions for online consent form, web-based survey (Nettskjema), and secure storage of research data (TSD).

We will collect directly identifiable data, probably the participants’ e-mail addresses. This will enable us to delete responses from participants who wish to withdraw from the study, to avoid sending messages to students who do not wish to take part, and to link different responses from the same respondent. The latter is of scientific importance as it allows monitoring of each participant over time, thus making this a cohort study.

We do not plan to transfer the collected data out of Norway. If we do, all data will be fully anonymized.

We will ensure highly secure data collection and data storage in collaboration with the data handler, USIT at the University of Oslo. Data management will be in accordance with GDPR-regulations. Our Data Protection Officer at the Norwegian Centre for Research Data, has assessed the project plan, and found it satisfactory (19 August 2020).

We see few ethical dilemmas related to this project, apart from the need to ensure secure handling of the collected person identifiable data. The only burden for the participants concerns the time spent completing the survey. The Regional Ethical Committee has assessed and approved the project plan (24 August, REK s0r-0st A, reference number 172155).

### Dissemination of findings

We will likely publish the findings in the format of scientific article in an open access medical journal.

### Power analysis and sample size

We have carried out a power analysis to estimate the necessary sample size needed to detect differences between students who are offered in-person instruction and students who are offered online instruction. We make a number of assumptions in estimating the necessary sample size.

We assume an incidence of COVID-19 of 0.23 % over a 10-week intervention period for students who are assigned to online instruction. This corresponds to the current level of disease in the age group 20-29 in Norway. If we wish to detect effects of in-person instruction that doubles the risk of disease, we would need 21,000 respondents to be 80% certain to detect an effect at 5% significance level. With a 50% response rate, we would need 42,000 students to be invited to the study. Oslo Metropolitan University alone has around 20,000 students.

Several of these assumptions can be challenged. It is highly uncertain whether the spread of COVID-19 will remain at its current level in the Norwegian student population, especially since the incidence has been rising for the past four weeks. Furthermore, there are few if any other studies of COVID-19 interventions in university settings, and thus it is difficult to estimate a likely effect size. Finally, the number of students who will be invited to this study remains to be seen, as it depends on the number of universities that agree to participate in the study.

As there is a large information need on the impacts of COVID-19 interventions and a scarcity of relevant studies, we furthermore believe the study can be beneficial even if underpowered and since a randomized assignment of participants was not possible. It may provide a solid foundation for more rigid, further intervention studies, and also possibly provide some evidence for the plausible range of effect sizes to be expected from moving to online instruction. A further benefit of the study is to map other important consequences of an online instruction intervention, such as effects on teaching satisfaction and life satisfaction.

We have submitted a registration form for the study to ClinicalTrials.gov.

## Data Availability

We plan to make the full anonymised data set publicly available.

## Author contributions

AF conceived the idea and wrote the first draft of the protocol. GJ and SH contributed in designing the study in the earliest phase. KT wrote the data analysis plan with substantial input from MF and AS. BL, IHS, AH, MF, AS and SSVW played key roles in developing the questionnaire. All authors read, commented on, and approved the final study protocol

## Acknowledgements

We thank Ole Petter Ottersen (Karolinska Institute) for contributing to the development of the idea behind this study. We also thank Per Martin Norheim-Martinsen (Oslo Metropolitan University) for supporting the project team in the development phase. Ann Oldervoll translated the questionnaire and the consent form to English, and Helen Ghebremedhin translated the questionnaire back to Norwegian for quality control (both Norwegian Institute of Public Health).

## Funding

No external funding received. The plan is that Oslo Metropolitan University and Norwegian Institute of Public Health will share the running costs.

## ATTACHMENT 1 – Questionnaire

**How old are you?**

Select 19 or younger

Etc.

50 or older

<u>Next page</u>

**With which gender do you identify?**

Woman

Man

Other

Prefer not to answer

<u>Next page</u>

Where were you born?

Norway

Europe (country other than Norway)

Africa

Asia

Australia/Oceania

North America

South or Central America

<u>Next page</u>

**Where were your parents/guardians born?**

(Select all that apply)

Norway

Europe (country other than Norway)

Africa

Asia

Australia/Oceania

North America

South or Central America

<u>Next page</u>

**What is your parent’s/guardian’s highest level of completed education?**

Primary/elementary school () Secondary or high school () College/university ()

Parent/guardian 1

Parent/guardian

<u>Next page</u>

**Have you been tested for the corona virus previously this year, before the beginning of the fall semester 2020?**

Yes

No

<u>Next page</u>

**In the past 14 days, have you been tested for the corona virus?**

Yes [the Norwegian version used No then yes this time, but the responses should always be in the same order so I am writing yes then no!]

No

<u>Next page</u>

**To what extent are you worried about being infected by the corona virus?**

To a very little extent [Or perhaps it would be better: not at all, slightly, somewhat, moderately, extremely]

To a small extent

To a moderate extent

To a great extent

To a very great extent

<u>Next page</u>

**In the past 14 days, have you been to a social gathering where you would guess that there were 20 or more people?**

Yes [Yes first again]

No

Do not know

<u>Next page</u>

**How many times have you participated in-person at a social gathering with your “faddergruppe” (buddy group) this fall semester 2020?**

None

1 time

2-4 times

5 times or more

<u>Next page</u>

**In the past 14 days, how many times have you drunk 6 or more alcoholic drinks?**

None

1 time

2 times

3 times

4 times

5 times or more Next page

**Which faculty are you enrolled in?**

Faculty of Health Sciences

Faculty of Education and International Studies

Faculty of Social Sciences

Faculty of Technology, Art and Design

Next page [Example, if respondent seletect Faculty of Technology, Art and Design on previous page]

**Which department are you affiliated with?**

Department of Civil Engineering and Energy Technology

Department of Mechanical, Electronic and Chemical Engineering

Department of Computer Science

Department of Product Design

Department of Art, Design and Dram

Other affiliation

<u>Next page</u>

**For which academic year are you taking most of your courses this semester?**

1^st^ year bachelor’s or 1^st^ year integrated master’s

2^nd^ year bachelor’s or 2^nd^ year integrated master’s

3^rd^ year bachelor’s or 3^rd^ year integrated master’s

1^st^ year master’s or 4^th^ year integrated master’s

2^nd^ year master’s or 5^th^ year integrated master’s

Other

I am not studying this semester Next page

**Overall, how satisfied have you been with the teaching you have received in the past 14 days?**

Scale from 0 to 10, where “0” is not satisfied at all and “10” is very satisfied

0 (not satisfied at all)

…

5 (neither satisfied nor dissatisfied)

…

10 (very satisfied)

Do not know

<u>Next page</u>

**In the past 14 days, approximately how many days have you been offered in-person instruction (on-campus or elsewhere)?**

Fill in the number of days

**In the past 14 days, approximately how many days have you been offered digital (online) instruction?** Fill in the number of days ______

<u>Next page</u>

**In the past 14 days, approximately how many days have you spent on campus (classes, self-study, colloquia and social)?**

Fill in the number of days_____

Have you been able to choose for yourself whether you wanted in-person or digital instruction?

Yes, to a large extent

Yes, to a small extent

No

Do not know

<u>Next page</u>

**In the past 14 days, how many days of off-campus work placement have you been offered?**

Fill in the number of days_____

**How many days have you had of off-campus work placement during the past 14 days?**

Fill in the number of days _____

<u>Next page</u>

**Approximately how many hours do you work (at a paying job) while studying per week?**

Fill in the number of hours ______

<u>Next page</u>

**Who do you live with now?**

Select all that apply

I live alone

With a romantic partner/spouse

With friend(s)/roommate(s)

With parents/guardians

With children

<u>Next page</u>

Who owns the housing unit you live in?

The welfare organization for students (SiO)

Professional private landlord Other private landlord My parents/guardians/relatives I/we own it Other

<u>Next page</u>

**How many other people do you live with? _____**

<u>Next page</u>

Approximately how many times have you travelled by public transportation (bus, tram, train, boat or plane) in the past 14 days?

<u>Next page</u>

**Overall, how satisfied are you with life right now?**

Scale from 0 to 10, where “0” is not satisfied at all and “10” is very satisfied

0 (not satisfied at all)

…

5 (neither satisfied nor dissatisfied)

…

10 (very satisfied)

Do not know

Send

Student survey on corona virus

Would you like to receive a receipt via email?

## ATTACHMENT 2 - Variables from the Common Student System (FS)

1. Sex (M, F)
2. Age
3. Basis of admission
4. Study progression: Average ECTS-credits the last 2 or 3 semesters
5. Grade Point Average (raw)
6. Year of secondary diploma
7. Grade Point Average (“school points”)
8. Grade Point Average (“admission points”)
9. Study programme code
10. Study programme name
11. Study level code
12. Field of study code
13. Field of study name
14. Share of full-time studies
15. Share professional training
16. NUS code
17. Students in study programme
18. Year of study
19. Course name (spring and autumn term 2020)
20. Course code (spring and autumn term 2020)
21. Course level (spring and autumn term 2020)
22. Course completion (spring and autumn term 2020)
23. Planned ECTS-credits (spring and autumn term 2020)
24. Outgoing exchange student (spring and autumn term 2020)
25. Incoming exchange student (spring and autumn term 2020)
26. Course grade (spring and autumn term 2020)
27. Average grade, all courses at institution
28. Institution number (NSD)
29. Institution number (FS)
30. Institution name
31. Place of study code DBH (NSD)
32. Place of study name
33. Campus
34. Faculty name

## ATTACHMENT 3 – Data analysis plan

A main concern in using a non-random design is that the treatment and comparison groups vary in observable and unobservable characteristics predictive of the outcome (infection). The main measure that safeguards against the comparison group holding more characteristics predictive of the outcome than the treatment group, is that the treatment (in-person instruction) was prescribed to groups of students, i.e. students did not themselves individually select in-person or online instruction. Whole classes where offered in-person instruction, and our main estimator is thus the impact on infection of in-person instruction for everyone offered in-person instruction (intention to treat, ITT [4]), not only those actually receiving in-person instruction.

It is still possible, of course, that the students being offered in-person instructions are more (or less) prone to become infected than the students referred to online instructions, in which case the estimates cannot be given a causal interpretation. The same will hold if the students who respond to the questionnaire (compared to those note responding) are more (or less) prone to infection *and* non-responding students are more prevalent in the group of students who are offered in-person instruction than in the group referred to online instructions (or v.v.). To explore this empirically, we will collect information that allows us to test the extent to which characteristics of the students differed in the group offered in-person instruction vs. online instruction at the outset. We will undertake balancing tests illustrated as follows:

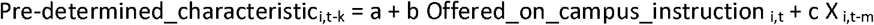

where *i* represents individual and *t* is time, k≥m≥0. Even if balanced, we would expect about 1 in 20 tests to be significant at the 5% level of significance, and if more tests are significant this strongly suggest that estimates should not be given a causal interpretation. If fewer are significant, we can still not conclude that the intervention is “as good as randomly” distributed and we should be very careful in making causal inference (1).

The most important pre-determined characteristic is likely the outcome variable before the intervention, i.e. already having been infected. We will collect data on previous infection as well as other (secondary) pre-determined outcome variables and characteristics that may be correlated with infection. These are age, gender, birthplace, parents’ birth place, socio economic status (parents’ education level), GPA points at admission, admission basis, exam results previous semester (where applicable), planned study credits, social behavior, living conditions, and use of public transport.

Importantly, we know *a priori* that some pre-determined variables are not balanced, by construction of the intervention. Most clearly, it was a stated goal for many universities that first-year students should be offered more in-person instruction than more senior students. Also, some fields of study were more dependent on in-person instruction than others, and were given priority. All balancing tests will therefore be performed with and without controlling for these “pre-stratification” variables (the vector X). Specifically, we will consider including the following

- Dummies for year of study program
- Dummies for field of study
- Dummies for courses
- Age and gender

We will also consider models where two or all these variables are interacted. Interactions will reduce the power to detect imbalance, but since the point is to run the exact same regression on the actual outcome variable, including many interactions will also reduce the power to detect effects. This isthus a classical trade-off between consistency and power. Since each university made different considerations, in what groups of students where offered more in-person instruction, these “pre-stratification” variables will be adapted to the policy, as described by each university before the intervention took place (or was changed).

Our population parameter (latent) of main interest is the effect of the intervention (offering in-person instruction) on the outcome (e.g. disease among the students). This ITT effect may be attempted estimated in a model illustrated as follows:

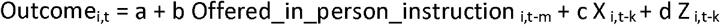

where k≥m≥0. X will include the same “pre-stratification” variables as in the balance test. With the sole purpose to improve precision, we will also consider including more pre-determined variables (i.e. the vector Z, like sex, age, place of birth, socioeconomic background, given that these variables are not included in X). When assessing learning outcomes, GPA points at admission (by cohort and admission basis) will be included as control variables.

The treatment variable will be operationalized both as two dummy variables: 80% or more of offered instruction time (excluding practice-time) is in-person vs. less than 80%; from 75^th^ percentile upward with in-person instruction vs. from 25^th^ downward with in-person instruction; and as a continuous variable, i.e. percent of teaching time offered in-person. Because the virus may spread more and more as the campus becomes more and more crowded, we will also estimate models allowing increasing marginal effects on disease as the percent with in-person instruction increases (several dummies).

Our primary outcome is (self-reported) infection, and secondary outcomes are

- COVID-19 test taken (dummy, logistic regression)
- Teaching satisfaction (scale, linear regression or multinomial logistic regression)
- Quality of life (scale, linear regression or multinomial logistic regression)
- Learning outcomes (completion and exam performance, linear regression or multinomial logistic regression)

In addition to ITT, we will also estimate the following “endogenous” model (using same estimation methods as described above):

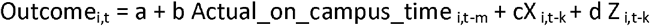

Moreover, we will estimate the local average treatment effect (LATE) in the following two stage model [4]

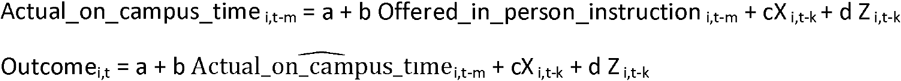

Where 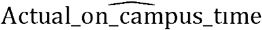 in the second equation can be considered as the estimate from the first stage (using e.g. 2SLS (1)). LATE is thus providing the effect of actually participating on campus for the students being moved from online to in-person by the offer. There are several strong assumptions necessary to give this a causal interpretation, where the exclusion restriction is typically considered the most important in practice: There is no conditional (on X and Z) direct effect of the offer of in-person instruction on the outcome, except through actual on-campus time, i.e. that the offer can be excluded from the second equation. The condition will be violated, for example, if an offer also affects transmission of the virus through more students meeting in other settings than on-campus (like more private parties). This assumption may be hard to defend, and the ITT estimate may be more reliable and relevant for decisions to be taken by the universities in balancing in-person and online instructions (1).

## Notes

### Competing Interest Statement

The authors have declared no competing interest.

### Clinical Trial

NCT04529421

### Funding Statement

The study is being funded by Oslo Metropolitan University and Norwegian Institute of Public Health. No external funding is foreseen.

### Author Declarations

Regional Ethics Committee, South-East PO BOX 1130, Blindern N-0318 Oslo Norway

